# Evaluating Large Language Model Diagnostic Performance on JAMA Clinical Challenges via a Multi-Agent Conversational Framework

**DOI:** 10.1101/2025.08.20.25334087

**Authors:** Karl L. Sangwon, Jeff Zhang, Robert Steele, Jaden Stryker, Jin Vivian Lee, Joanne Choi, Krithik Vishwanath, Daniel Alexander Alber, Douglas Kondziolka, Michal Mankowski, Eric Karl Oermann

**Author notes:** **Corresponding author during review** Karl L. Sangwon, NYU Langone Department of Neurosurgery, 550 First Ave, New York, NY 10016. **Corresponding author post-review** Eric K. Oermann, NYU Langone Department of Neurosurgery, 550 First Ave, New York, NY 10016. **Disclosures:** EKO reports employment in Eikon Therapeutics; equity in Artisight Inc., Delvi Inc., MarchAI Inc.; consulting for Sofinnova, Google. **Disclosure of Funding:** None. Author Contributions: Karl L. Sangwon: Study and Engineering Design, Manuscript Writing, Statistical Analysis; Jeff Zhang: Engineering Assistance with Program Library Development; Robert Steele: Engineering Assistance with High-Performance Computing Environment Setup; Jaden Stryker: Engineering Assistance with Language Model Inference Setup; Jin Vivian Lee, Joanne Choi: Assist with Experiments, Manuscript Edits; Krithik Vishwanath: JAMA question obtainment; Daniel Alexander Alber: Manuscript Edit; Douglas Kondziolka: Administrative Support; Michal Mankowski: Administrative Support; Eric K. Oermann: Study Design, Administrative Support, Study Supervision.

## Abstract

**Background & Objective:** Standard clinical LLM benchmarks use multiple-choice vignettes that present all information up front, unlike real encounters where clinicians iteratively elicit histories and objective data. We hypothesized that such formats inflate LLM performance and mask weaknesses in diagnostic reasoning. We developed and evaluated a multi-AI agent conversational framework that converts JAMA Clinical Challenge cases into multi-turn dialogues, and assessed its impact on diagnostic accuracy across frontier LLMs.

**Methods:** We adapted 815 diagnostic cases from 1,519 JAMA Clinical Challenges into two formats: (1) original vignette and (2) multi-agent conversation with a Patient AI (subjective history) and a System AI (objective data: exam, labs, imaging). A Clinical LLM queried these agents and produced a final diagnosis. Models tested were O1 (OpenAI), GPT-4o (OpenAI), LLaMA-3-70B (Meta), and Deepseek-R1-distill-LLaMA3-70B (Deepseek), each in multiple-choice and free-response modes. Free-response grading used a separate GPT-4o judge for diagnostic equivalence. Accuracy (Wilson 95% CIs) and conversation lengths were compared using two-tailed tests.

**Results:** Accuracy decreased for all models when moving from vignettes to conversations and from multiple-choice to free-response (p<0.0001 for all pairwise comparisons). In vignette multiple-choice, accuracy was O1 79.8% (95% CI, 76.9%–82.4%), GPT-4o 74.5% (71.4%–77.4%), LLaMA-3 70.9% (69.5%–72.2%), Deepseek-R1 69.0% (67.5%–70.4%). In conversation multiple-choice: O1 69.1% (65.8%–72.2%), GPT-4o 51.3% (49.8%–52.8%), LLaMA-3 49.7% (48.2%–51.3%), Deepseek-R1 34.0% (32.6%–35.5%). In conversation free-response: O1 31.7% (28.6%–34.9%), GPT-4o 20.7% (19.5%–22.0%), LLaMA-3 22.9% (21.6%–24.2%), Deepseek-R1 9.3% (8.4%–10.2%). O1 generally required fewer conversational turns than GPT-4o, suggesting more efficient multi-turn reasoning.

**Conclusions:** Converting vignettes into multi-agent, multi-turn dialogues reveals substantial performance drops across leading LLMs, indicating that static multiple-choice benchmarks overestimate clinical reasoning competence. Our open-source framework offers a more rigorous and discriminative evaluation and a realistic substrate for educational use, enabling assessment of iterative information-gathering and synthesis that better reflects clinical practice.

---

Standard clinical LLM benchmarks rely on multiple-choice vignettes where all relevant information is presented upfront—failing to reflect how clinicians iteratively gather and synthesize data in real encounters.^1-4^ We hypothesized that this format may inflate LLM performance and mask weaknesses in diagnostic reasoning. To address this, we developed and evaluated a multi-AI agent conversational framework that converts *JAMA Clinical Challenge* case vignettes into dynamic multi-turn dialogues simulating a more realistic diagnostic process. We assessed the effect of this conversational format on diagnostic accuracy across frontier LLMs.

## Methods

We adapted 815 diagnostic cases from 1,519 publicly available *JAMA Clinical Challenges* into two formats: (1) original vignette and (2) multi-agent conversation involving Patient AI (subjective history) and System AI (objective clinical findings: exam, labs, imaging). A Clinical LLM interacts with these agents and synthesizes information to generate a diagnosis (**Figure 1**). Prompt details are available in **eTable** (**Supplement 1)**.

**Figure 1.**
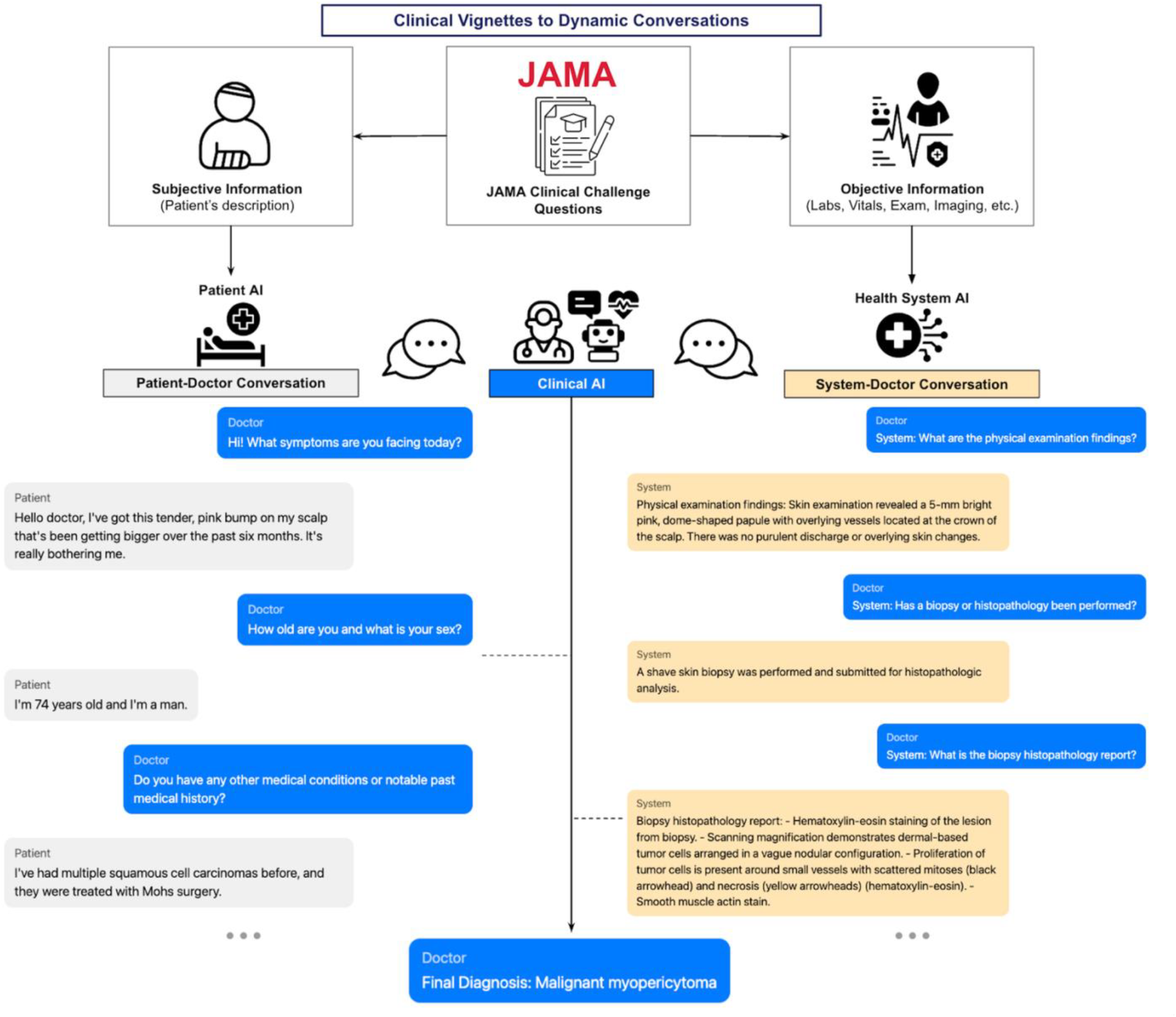
Multi-AI Agent Framework for Converting Clinical Vignette Questions to Conversations. The framework transforms JAMA Clinical Challenge questions into dynamic conversations by separating information into subjective (patient-reported) and objective (clinical data) components. A Clinical AI or doctor navigates two distinct conversational interactions: (1) Patient-Doctor Conversation through a Patient AI that provides subjective information based on the patient’s description, and (2) System-Doctor Conversation through a Health System AI that supplies objective clinical data (laboratory results, vital signs, examination findings, and imaging). The Clinical AI or human user integrates information from both channels to formulate a final diagnosis. Example conversation snippets demonstrate the natural flow of history-taking and clinical data gathering through these parallel interfaces.

We evaluated GPT-4o (OpenAI), LLaMA-3-70B (Meta), Deepseek-R1-distill-LLaMA3-70B (Deepseek), and O1 (OpenAI). Deepseek and O1 models were selected for their reported multi-step reasoning capabilities—Deepseek via training on mathematical problem-solving tasks, and O1 as a reasoning-optimized foundational model. Each model and case were tested in both multiple-choice and free-response formats. A separate GPT-4o model judged free-response answers for diagnostic equivalence. Conversations were checked for clinical coherence by medical trainees and attending physicians. Performance metrics included diagnostic accuracy and average conversation length. Accuracy was reported with 95% confidence intervals using Wilson’s methods and compared using 2-tailed t tests.

## Results

O1 achieved the highest accuracy across all formats, followed by GPT4o, LlaMA-3 and Deepseek-R1. All models had lower accuracy in conversational formats vs vignettes, and in free-response vs multiple-choice formats (p<.0001, all pairwise). (**Figure 2A**)

**Figure 2.**
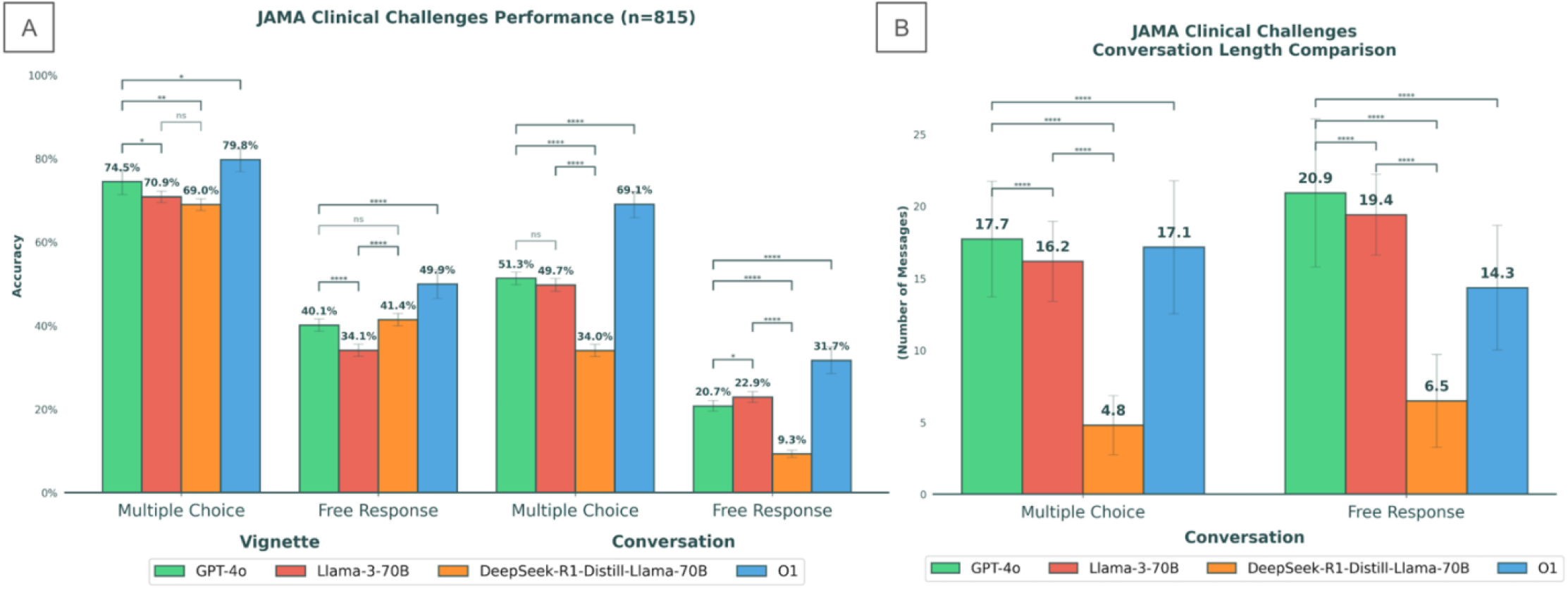
LLM Performance Analysis Across Assessment Formats. (A) Diagnostic accuracy of four different LLMs on JAMA Clinical Challenge questions across different formats. Performance dropped significantly when multiple choices were removed, and when converted into conversation format where models had to interact with other AI models to iteratively collect information for diagnosis (p<0.0001 for each). Error bars represent 95% confidence intervals. (B) Analysis of LLM conversation lengths across question formats. The average number of conversational exchanges for multiple choice was lower than free response formats. Error bars represent one standard deviation. **** denotes p<0.0001, *** denotes p<0.001, ** denotes p<0.01, * denotes p<0.05, ns denotes non-significant p-value.

In vignette multiple-choice, accuracy was: O1, 79.8% (95%CI, 76.9%-82.4%); GPT-4o, 74.5% (71.4%-77.4%); LLaMA-3, 70.9% (69.5%-72.2%); Deepseek-R1, 69.0% (67.5%-70.4%). In vignette free-response: O1, 49.9% (46.5%-53.4%); GPT-4o, 40.1% (38.6%-41.6%); LLaMA-3, 41.4% (32.6%-35.6%); Deepseek-R1, 34.1% (39.9%-42.9%) (p<.0001, all pairwise).

In conversation multiple-choice, O1 again led at 69.1% (65.8%-72.2%), followed by GPT-4o (51.3%, 49.8%-52.8%), LLaMA-3 (49.7%, 48.2%-51.3%), and Deepseek-R1 (34.0%, 32.6%-35.5%). In conversation free-response: O1, 31.7% (28.6%-34.9%); GPT-4o, 20.7% (19.5%-22.0%); LLaMA-3, 22.9% (21.6%-24.2%); Deepseek-R1, 9.3% (8.4%-10.2%) (p<.0001, all pairwise).

In conversation multiple-choice, GPT-4o had the longest interactions (17.7 messages), followed by O1 (17.1), LLaMA-3 (16.2), and Deepseek-R1 (4.8). In conversation free-response, GPT-4o again led (20.9), followed by LLaMA-3 (19.4), O1 (14.3), and Deepseek-R1 (6.5) (**Figure 2B**).

## Discussion

Our study suggests that static clinical vignettes overestimate LLM’s diagnostic reasoning capabilities. By simulating more realistic clinical encounters through a multi-agent, multi-turn conversational framework applied to *JAMA Clinical Challenge* cases, we observed substantial performance drops across models—particularly in free-response formats—underscoring challenges in iterative information gathering and synthesis.

Deepseek-R1-distilled-Llama-3, a model trained for mathematical problem-solving reasoning, performed worst in free-response conversations—often failing to ask appropriate follow-up questions or retrieve key clinical details. This suggests that pretraining optimized for structured, single-turn tasks may limit multi-turn diagnostic reasoning.^4^ In contrast, O1 consistently outperformed others across all formats and required fewer turns, suggesting more efficient and conversational reasoning.

These findings indicate that traditional multiple-choice benchmarks may overstate LLMs’ clinical competence.^5,6^ Conversational evaluations offer a more rigorous and discriminative approach, capable of distinguishing between models with and without advanced multi-turn reasoning capabilities. Our open-source framework provides a scalable, reproducible method for evaluating LLMs in more clinically reflective settings.

Limitations include reliance on static vignettes as the information source and the absence of human comparator data. Future studies would benefit from expanded source data and evaluation of clinician-AI interactions using this framework for educational and diagnostic applications, particularly in domains requiring iterative clinical reasoning such as differential diagnosis or triage.

## Data Availability

All data produced are available online at https://jamanetwork.com/collections/44038/clinical-challenge

https://jamanetwork.com/collections/44038/clinical-challenge

